# Progress towards the End TB Goals in South Africa: A comparative cost-effectiveness analysis of tuberculosis interventions

**DOI:** 10.64898/2026.01.26.26344817

**Authors:** Mmamapudi Kubjane, Lise Jamieson, Leigh F. Johnson, Kamban Hirasen, Lezanie Coetzee, Clive Ramushu, Denise Evans, Pren Naidoo, Gesine Meyer-Rath

## Abstract

**Background:** South Africa’s National Tuberculosis (TB) Programme aims to achieve targets set by the World Health Organization’s End TB Strategy, including an 80% reduction in TB incidence and a 90% reduction in TB mortality by 2030, compared to 2015 levels. We were tasked to evaluate 1) the impact on TB incidence and mortality of scaling up individual interventions focusing on TB preventive therapy, screening, testing and linkage to treatment as included in the 2023–2028 National Strategic Plan for TB (NSP); 2) the budget required for implementing the NSP; 3) the cost-effectiveness of scaling up individual interventions aligning with NSP targets, and 4) the health impact and cost-effectiveness of additional aggressive screening scenarios to reach the End TB targets.

**Methods:** We used the Thembisa TB model and public-sector costs to estimate the incremental cost-effectiveness per life year saved (LYS) for individual and combined interventions under various scenarios from 2023 to 2042. The NSP scenario included expansions of TB preventive therapy (TPT), symptom screening at primary health clinics and testing with Xpert, screening of household contacts of people with TB, and community-based screening (door-to-door and digital chest X-ray), as well as targeted universal testing for TB (TUTT) in people living with HIV (PLHIV), household contacts of people with TB and individuals with a history of TB, and reduction of initial loss-to-follow-up (ILTFU), all to levels deemed feasible by NSP stakeholders. We also assessed two hypothetical scenarios that largely maximise screening: Max scenario 1 (TPT for PLHIV, quarterly symptom screening for all adults) and Max scenario 2 (TPT for PLHIV, with eligible adults receiving yearly chest X-rays, TB testing, and TPT for household contacts).

**Results:** The NSP achieved a 44% reduction in TB incidence and a 55% reduction in mortality by 2030 relative to 2015, while the aggressive scenarios (Max 1 and Max 2) achieved 57% and 56% reductions in incidence, respectively; and 75% and 71% reductions in mortality, respectively. Among individual NSP interventions, over the period 2023-2043, TPT for PLHIV and Xpert testing for symptomatic individuals seeking TB care were cost-saving. Symptom screening for household contacts ($12/LYS), ILTFU reduction ($13/LYS), TUTT for household contacts ($84/LYS), and TPT for household contacts ($106/LYS) were the most cost-effective interventions. Compared to baseline, the NSP scenario increased cost by 57%, saving 6.6 million life-years at $308/LYS over the period 2023-2042. The Max 1 and Max 2 scenarios increased the cost significantly, by 329% and 1526%, respectively. Max 1 saved 16.5 million life-years at $712/LYS, while Max 2 saved 14.6 million life-years at $3,774/LYS.

**Conclusion:** Scaling up TB interventions to NSP coverage targets will substantially save lives and reduce TB morbidity. However, the End TB targets will not be met even under additional scenarios considered, including more aggressive prevention and screening. Accelerating progress would require both improving efficiencies of existing strategies and developing new tools including low-cost diagnostics and effective TB vaccines.

## Background

South Africa has high rates of drug-susceptible (DS) tuberculosis (TB), drug-resistant TB (DR-TB) and HIV-associated TB, being among the top 30 countries accounting for 86-90% of the global burden of these conditions [1]. In 2023, approximately 329,000 (95% confidence interval (CI): 310,000–346,000) South Africans developed TB, 13,000 (95% CI: 7,600–29,000) developed DR-TB, and 62,000 (95% CI: 56,000–67,000) died of TB [2,3]. The burden of TB is much higher among men, with the TB prevalence 1.6 times higher in men than women [4], driven by factors such as smoking, alcohol use and barriers to TB and HIV care [3]. HIV infection remains a major driving factor for TB in South Africa, with approximately 8 million individuals living with HIV [5,6] and approximately 60% and 66% of these individuals diagnosed with DS-TB and DR-TB being co-infected with HIV, respectively [1].

In addition to its impact on morbidity and mortality, TB also imposes a significant economic burden on affected individuals. Although treatment is free at the point of care for those without private insurance, the World Health Organization reports that 56.2% (55.9% for DS-TB and 64% for DR-TB) of South African TB patients experience TB-related expenses that exceed 20% of their annual household income [1]. Moreover, the national TB programme faces substantial costs in providing TB-related care services. The National AIDS Spending Assessment Plus (NASA+) report indicated that in 2019/2020, TB-related expenses reached $206 million [7]. Most of this expenditure (62%) was allocated to treating DR-TB, followed by treating DS-TB (20%), screening and diagnostic interventions (16%) and prevention (2%) [7].

Progress has been made in mitigating the impact of TB on health in South Africa. Between 2009 and 2019, new TB cases decreased by approximately 30%, and TB-related mortality declined by about 48%, with large proportions of these declines being driven by the provision of antiretroviral treatment (ART) for HIV and intensified TB screening efforts [8]. However, the COVID-19 pandemic has set back these efforts due to reduced testing rates and treatment initiation [9–11]. To achieve the ambitious End TB targets of reductions in TB incidence by 80% and TB deaths by 90% relative to 2015 levels, substantial financial support for the National TB Programme is crucial. Additionally, funds need to be allocated efficiently across interventions.

Mathematical models facilitate budget planning, guide resource allocation, and maximise epidemiological impact. Past TB mathematical modelling analyses have assessed the cost and effectiveness of scaling-up TB prevention, case-finding strategies, and improving treatment while also accounting for resource constraints [12–14], and the allocative efficiency of TB interventions in a single province [15]. The first South African HIV/TB Investment Case, conducted in 2016, was used to support policy changes and budget expansions for implementing recommended interventions such as TB preventive therapy (TPT) for people living with HIV (PLHIV), TB symptom screening at healthcare facilities, scaled-up Gene Xpert testing, and ensuring successful treatment [16,17].

While these previous TB modelling studies have provided valuable insights on the impacts of intervention health and economic impacts, most addressed specific research questions rather than ongoing policy requirements. These analyses used simplified HIV components or external HIV estimates rather than dynamically simulating HIV epidemic and intervention effects on TB. Additionally, these studies largely use WHO estimates [1] – themselves model outputs with substantial uncertainty – rather than South Africa’s local data (TB testing and diagnoses, recorded deaths, routine programme data). This study addresses these gaps by using a comprehensively integrated TB-HIV model calibrated to local data and public sector TB programme costs to provide cost-effectiveness estimates for ongoing policy guidance on resource allocation.

We were tasked by the South African National TB Programme to update the 2016 TB Investment Case to evaluate: 1) the impact on TB incidence and mortality of scaling up individual interventions included in the 2023–2028 National Strategic Plan for TB (NSP); 2) the budget required for implementing the NSP; 3) the cost-effectiveness of scaling up individual interventions aligning with NSP targets, and 4) the health impact and cost-effectiveness of additional aggressive screening scenarios to reach the End TB targets.

Building on the 2016 methodology [16,17], we assessed the health impact, cost, and cost-effectiveness of scaling up existing and new TB interventions using a TB transmission model that is calibrated to current TB and HIV programme coverage. Our update incorporates recent advances such as shorter DR-TB treatment regimens [18], improving linkage to treatment initiation, expanded preventive therapy [19], and innovative case-finding strategies - including targeted universal testing for TB (TUTT) in high-risk groups including PLHIV, individuals who have had a TB episode in the last two years[20,21], door-to-door symptom screening, and digital chest X-ray screening for asymptomatic adults [22].

## Methods

### The epidemiological model

We used the Thembisa TB transmission model, a deterministic compartmental model developed to simulate the adult DS-TB and DR-TB epidemic in South Africa, to assess the impact of several TB interventions [8]. The model has been calibrated to South African data, including sex-stratified recorded numbers of TB deaths from the country’s vital registration system [23], numbers of people initiating treatment, deaths on treatment, HIV prevalence in persons treated for TB treatment [24], the number of microbiological tests performed [25] and TB prevalence data [26]. The model was recently extended to include DR-TB [27] and was calibrated to national DR-TB prevalence survey data and to numbers of patients diagnosed with rifampicin-resistant TB and initiating second-line treatment from the Electronic Drug-Resistant TB register (EDRweb) [28]. To dynamically incorporate the effects of HIV and ART on TB, both DS-TB and DR-TB are integrated into the Thembisa HIV model that simulates the South African HIV epidemic and addresses policy questions relating to HIV prevention and treatment [29]. The Thembisa TB and HIV models, as well as the DR-TB extensions, have been described previously elsewhere [2,5,8,27]. We used the mean parameter values of the posterior estimates obtained from the latest Thembisa TB and HIV model calibration [2,30].

### Baseline interventions and new interventions, coverage levels, target populations and scenarios

The baseline scenario for the current TB programme includes the following interventions: preventive therapy for PLHIV (six months of isoniazid preventive therapy (IPT) and three months of isoniazid plus rifapentine (3HP)). TB symptom screening in public health facilities is conducted, followed by Xpert testing for individuals with TB symptoms seeking TB care and those with TB-like symptoms attending health facilities for other reasons. Culture testing is carried out for PLHIV with a negative Xpert test result, and treatment for DS-TB and DR-TB is provided (Table 1). Newly suggested interventions, identified during extensive stakeholder engagement in preparation for the 2023-2027 National Strategic Plan for HIV, TB and STIs (NSP) (as indicated in Table 1), include screening household contacts of people with TB and community-based screening, such as door-to-door and digital chest X-rays; universal testing for specific groups such as PLHIV, household contacts of people with TB, and individuals with a previous TB episode, alongside linkage to treatment [31], such as shorter DR-TB treatment regimens, reduced from 24 months to 9 months [32].

**Table 1:** Coverage assumptions by intervention and scenario implemented annually.

| Description of intervention and target population | Baseline | National Strategic Plan | Max 1 | Max 2 |
| --- | --- | --- | --- | --- |
| **<u>Prevention</u>** |  |  |  |  |
| Proportion of eligible* PLHIV initiated on IPT or 3HP (converted from rates of monthly TPT uptake) | 30% | 48% | 60% | 60% |
| • % initiated on 3HP (other proportion initiated on IPT) | 50% | 90% | 100% | 100% |
| Proportion of eligible adult household contacts of TB patients (converted from rates of monthly TPT uptake) | 0% | 55% | 0% | 100% |
| • % initiated on 3HP (other proportion initiated on IPT) | 0% | 90% | 0% | 100% |
| Efficacy of 3HP in curing infection | 50% | 50% | 50% | 50% |
| **<u>Screening</u>** |  |  |  |  |
| Proportion of adult household contacts of index TB cases screened for symptoms | 0% | 87% | 0% | 0% |
| Proportion of adults screened for symptoms door-to-door (approximated from the average number of annual symptom screens) | 0% | 11% | 400% | 100% |
| Proportion of door-to-door screened offered dCXR if asymptomatic | 0% | 8% | 0% | 100% |
| **<u>Testing (passive)</u>** |  |  |  |  |
| Proportion of symptomatic TB patients tested with Xpert if attending health services for TB care | 16% | 20% | 16% | 16% |
| Proportion of individuals with TB-like symptoms tested with Xpert if attending health services for other reasons | 2% | 0% | 0% | 0% |
| Proportion of culture testing for PLHIV who test negative on Xpert | 30% | 35% | 30% | 30% |
| **<u>Testing (irrespective of TB symptoms)</u>** |  |  |  |  |
| Proportion TUTT for PLHIV on ART (approximated from the average annual number of TUTT for PLHIV on ART) | 0% | 51% | 0% | 0% |
| Proportion of TUTT for individuals with a history of TB (approximated from the average annual number of TUTT for those with a history of TB) | 0% | 12% | 0% | 0% |
| Proportion of household contacts tested irrespective of symptoms | 0% | 74% | 0% | 100% |
| **<u>Linkage to treatment</u>** |  |  |  |  |
| Proportion of individuals linking to care after diagnosis and starting on TB treatment | 82.4% | 90.3% | 90.3% | 90.3% |

Abbreviations: 3HP, 3 months of isoniazid-rifapentine; dCXR, digital chest X-ray; ILTFU, initial loss to follow-up; IPT, isoniazid preventive therapy (6 months); NA, not applicable; PLHIV, people living with HIV; TB, tuberculosis; TPT, tuberculosis preventive therapy; TUTT, targeted universal testing for tuberculosis. \*TPT eligible populations include total PLHIV on ART, HIV diagnosed, and those starting ART.

Amongst the novel interventions considered for the NSP, the door-to-door screening intervention assumes that any adult in the population will be screened at home for symptoms by community healthcare workers. The digital chest X-ray screening intervention is implemented as part of door-to-door symptom screening, in which individuals who are asymptomatic after a symptom screen are invited to undergo further screening with a mobile digital chest X-ray. The assumed sensitivity of digital chest X-ray is 83%, and the specificity is 63% [22]. Household contact screening targets persons who share a household with active TB patients, with the assumption that 68% of TB-diagnosed household contacts are then linked to TB treatment [33].

Targeted universal testing for TB (TUTT) is modelled as testing in people regardless of their TB symptom status, implemented at public health facilities [20,21] and targeting those who fall into one of three categories: people living with HIV on ART, those who self-report a history of a TB episode (everyone who ever had TB), and household contacts of patients with active TB. Lastly, we modelled a linkage-to-treatment intervention for patients diagnosed with TB who are not yet on treatment. This intervention reduces initial loss to follow-up (ILTFU) after TB diagnosis and increases treatment initiation rates by 42% according to the LINKEDin study [31]. We do not model paediatric TB dynamically; instead, we assume a constant child-to-adult TB incidence ratio based on WHO estimates [1]. We then apply this ratio to the Thembisa adult TB incidence to estimate the number of paediatric TB cases and to approximate the treatment need for this group, along with the treatment cost.

### Modelling target populations and intervention scenarios

We generated target populations for each intervention using the Thembisa TB model. In the baseline scenario, we maintained existing TB programme interventions without improvement. For the NSP scenario, intervention coverage levels (Table 1) were established through discussions with stakeholders giving input into the NSP, including National TB Programme staff from the National Department of Health and the TB Think Tank, a national network of TB experts providing evidence-based advice to the South African National Department of Health on TB control [34].

Additionally, we implemented two hypothetical aggressive intervention scenarios – Max Scenario 1 and Max Scenario 2 – for exploratory analysis. Max Scenario 1 includes quarterly door-to-door symptom screening and reduced loss-to-follow-up before treatment initiation. Max Scenario 2 assumes all adults are screened with a digital chest X-ray in a door-to-door context, TPT for household contacts, and TUTT of contacts (Table 1). These scenarios assess the model’s sensitivity to intensified community-based and targeted interventions and explore whether End TB targets are achievable under such conditions.

### Outcomes

We estimated the epidemiological impact of scaling up each intervention and scenario on the expected number of new TB cases, TB-related deaths, years of life lost (YLL) due to TB, and disability-adjusted life years (DALYs) associated with TB over 2023–2042. Years of life lost were estimated by multiplying age-specific model-estimated TB deaths by the expected remaining years of life at the age of death, based on the West level 26 life table [35]. Disability-adjusted life years were calculated as the sum of YLL and years lived with disability (YLD) due to TB. Years lived with disability were calculated by multiplying the Thembisa model estimated annual incident TB (for PLHIV and people without HIV) and duration of TB disease, and TB-related disability weights for PLHIV and people without HIV [36]. The TB disability weights (0.33 for people without HIV; and 0.408 for PLHIV) were as estimated by the Global Burden of Disease study [35,36]. Lastly, we calculated cumulative TB incidence and mortality in each scenario and the percentage reductions due to interventions compared to the baseline.

### Costs and cost-effectiveness

The cost inputs used in this analysis were obtained from the National TB Cost Model [37], which was informed by a systematic review of all available primary cost data for TB interventions in South Africa, augmented by bottom-up cost analyses from recent studies and ingredient-based cost analysis for novel interventions (Table 2). We analysed cost and resource utilization from the perspective of the provider, the South African government. Across interventions, we adjusted prices and salaries to the 2023/24 public-sector values. All costs were converted to United States Dollars (USD) using the average exchange rate of 1 USD to 18.45 South African Rand for July 2023–June 2024 [38]. Costs are presented undiscounted and not adjusted for inflation over the period modelled. We analysed cost-effectiveness over a 20-year time horizon (2023-2042), compared to a baseline of currently available TB interventions, estimating the incremental cost per new active TB case averted, per life year saved and per DALY averted.

**Table 2:** Input intervention costs for tuberculous prevention, screening, testing, linkage to treatment, and treatment (2023/2024 USO)

| Intervention category and description of interventions and target populations | Average cost | Unit |
| --- | --- | --- |
| **<u>Prevention</u>** |  |  |
| IPT for PLHIV on ART or household contacts of TB patients are eligible | 11.0 | per person initiated |
| 3HP for PLHIV on ART or household contacts of TB patients eligible | 22.8 | per person initiated |
| **<u>Screening</u>** |  |  |
| Symptom screening (passive) at public health facilities | 0.7 | per person screened |
| Symptom screening for household contacts per index TB case | 1.5 | per person screened |
| Community-based door-to-door symptom screening | 1.7 | per person screened |
| Community-based screening with digital chest X-ray (dCXR) for asymptomatic individuals | 41.1 | per person screened |
| **<u>Testing</u>** |  |  |
| Xpert testing (passive) at public health facilities | 16.8 | per patient tested |
| Culture testing for PLHIV who test negative on Xpert | 5.6 | per patient tested |
| TUTT for PLHIV | 16.8 | per patient tested |
| Linkage to treatment for PLHIV diagnosed with TB | 3.0 | per patient diagnosed |
| TUTT for individuals with a previous TB episode | 16.8 | per patient tested |
| Linkage to treatment for individuals with a previous TB episode diagnosed with TB | 3.0 | per patient diagnosed |
| TUTT for household contacts | 16.8 | per patient tested |
| Linkage to treatment for household contacts diagnosed with TB | 3.2 | per patient diagnosed |
| **<u>Linkage to treatment</u>** |  |  |
| Improved linkage to DS-TB treatment for individuals with a positive TB diagnosis | 1.7 | per patient diagnosed and not yet linked to treatment |
| **<u>Treatment</u>** |  |  |
| Drug-susceptible TB (outpatient) | 128.7 | per patient treated |
| Drug-susceptible TB (inpatient) | 298.9 | per patient admitted |
| Drug-resistant TB (inpatient and outpatient) | 5,287.9 | per patient treated |
| Treatment monitoring using smear microscopy | 4.7 | per patient treated |
| Paediatric TB treatment | 100.6 | per patient treated |

**Abbreviations:** 3HP, 3 months of isoniazid-rifapentine; ART, antiretroviral therapy; dCXR, digital chest X-ray; ILTFU, initial loss to follow-up; IPT, isoniazid preventive therapy (6 months); NA, not applicable; PHC, public healthcare clinic; PLHIV, people living with HIV; TB, tuberculosis; TPT, tuberculosis preventive therapy; TUTT, targeted universal testing for tuberculosis. TPT-eligible populations include all PLHIV on ART, those newly diagnosed with HIV, and individuals initiating ART.

### Sensitivity analysis

We conducted one-way sensitivity analyses varying intervention-specific unit costs by ±50% around base-case values in Table 2 (e.g., Xpert testing cost range: 8.41–25.23 USD), reporting changes in incremental costs and incremental cost per life year saved relative to the baseline while holding intervention coverages and effectiveness constant. Additionally, given the uncertainty in our assumption that 3HP has a 50% sterilising effect (i.e. clears infection), we varied this parameter at values: 10%, 25%, 50%, 75% and 90% efficacy, when provided to PLHIV on ART and household contacts of TB patients, and assessed the impacts on health outcomes life-years saved, DALYs and new TB cases averted, and incremental cost-effectiveness over the period 2023-2042.

## Results

### Tuberculosis incidence and mortality rates trends 2023-2042

In the baseline scenario, TB incidence and mortality rates were projected to decline gradually from 2015 to 2042 (Figure 1), and a sharp drop between 2020 and 2021 was largely due to social distancing and mask-wearing during the COVID-19 pandemic. This drop was followed by a rebound in 2022, partly due to reductions in social distancing and TB service interruptions. Projected reductions in TB incidence by 2030, relative to 2015, were 35% (baseline), 44% (NSP), 57% (Max 1), and 56% (Max 2) (Figure 1a), while reductions in mortality rates were 41% (baseline), 55% (NSP), 75% (Max 1), and 71% (Max 2) (Figure 1b).

**Figure 1:**
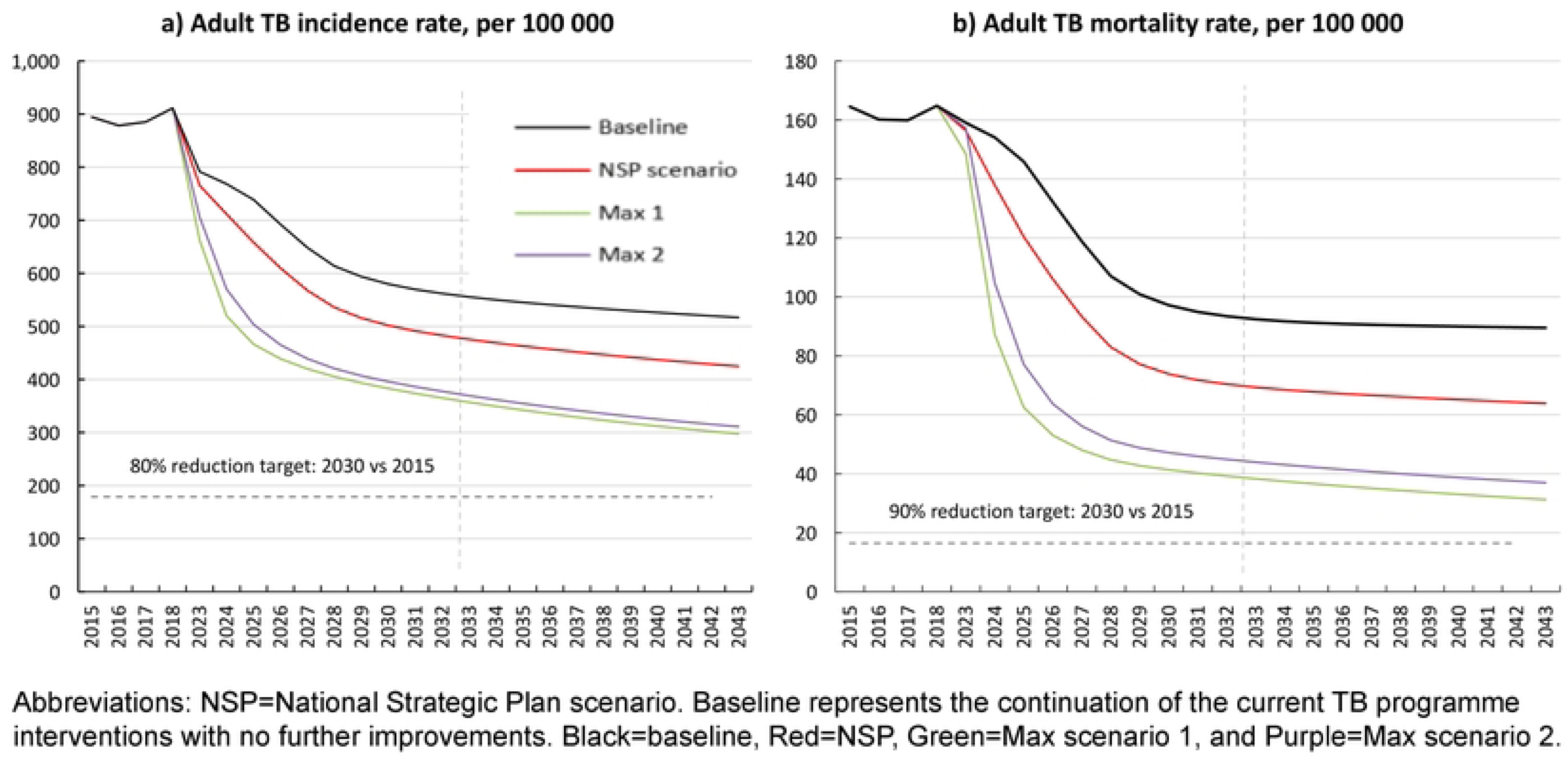
Tuberculosis incidence (a) and mortality (b) rates per 1C.

### Impact of interventions on incidence and mortality, cumulative reductions (2023-2042)

In the NSP scenario, the individual interventions with the greatest impact on TB incidence and mortality were increasing Xpert testing for symptomatic individuals presenting at public health facilities (7% reduction in incidence and 13% reduction in mortality), linking diagnosed TB patients to TB treatment (4% reduction in incidence and 7% in mortality); providing household contacts with TB preventive therapy (3% reduction in incidence and 4% in mortality) and testing household contacts irrespective of symptoms (2% reduction in incidence and 4% in mortality) (Figure 2). Door-to-door symptom screening would result in a 2% reduction in TB incidence, and adding digital chest X-ray for asymptomatic individuals would result in a further 0.3% reduction. The estimated mortality reductions from door-to-door symptom screening and digital chest X-ray for asymptomatic individuals were 3% and 0.2%, respectively.

**Figure 2:**
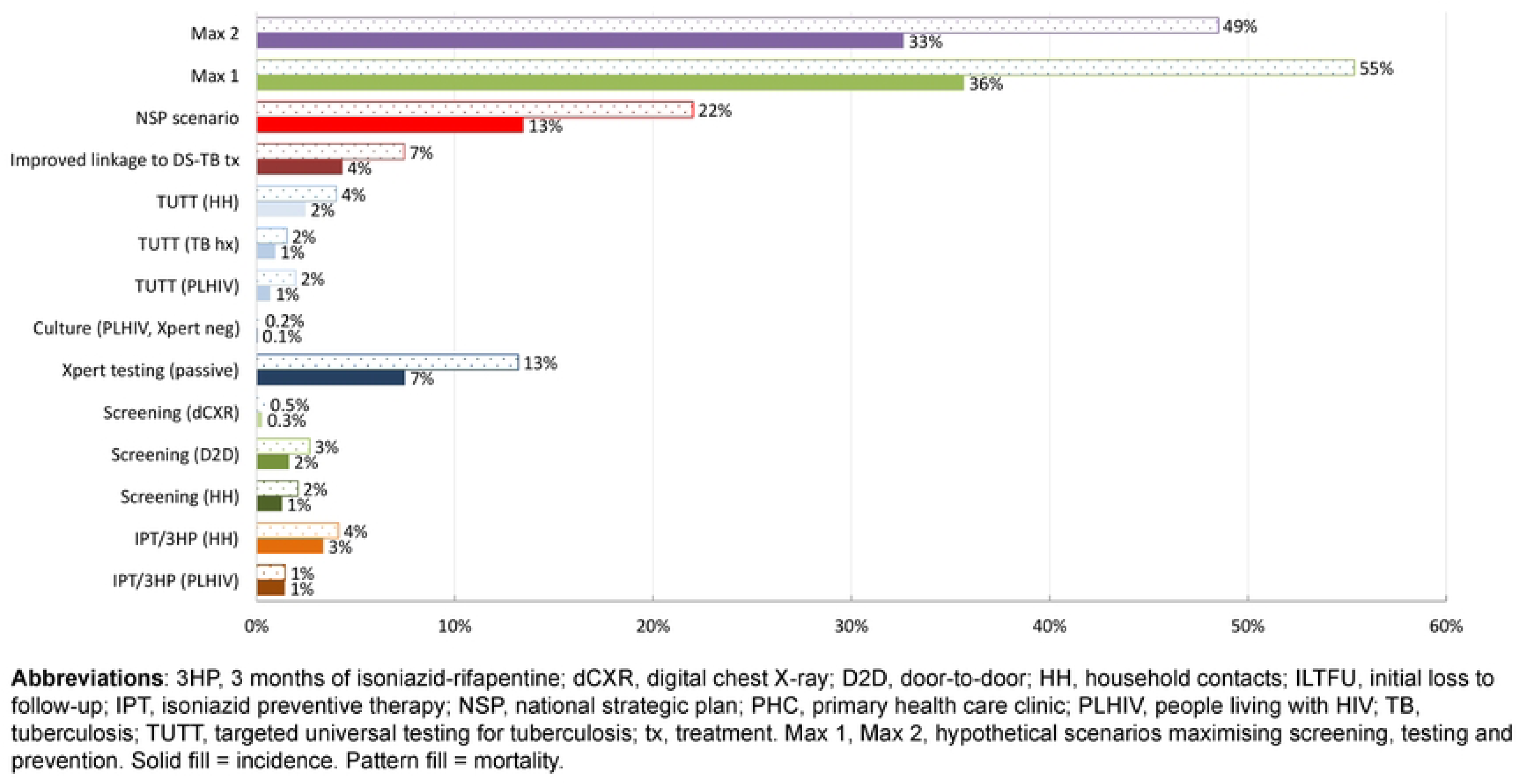
Projected percentage reduction in tuberculosis incidence.

### Budget required 2023-2027

Under the baseline scenario, the annual cost of the TB programme averaged $174 million over 2023-2027 (Figure 2a). The NSP scenario increased the annual total cost by 18%, from $242 million in 2023 to $284 million in 2027 (Figure 2a). The total budget over 2023-2027 was $1,282, a 48% increase over the baseline total budget (2023-2027), with diagnostic interventions and treatment constituting the greatest proportion of total cost. Xpert testing accounted for the greatest share of the budget (18% for passive, and 16% for TUTT for PLHIV), followed by DS-TB treatment (18%) and DR-TB treatment (16%).

In Max scenario 1, the estimated total budget for 2023-2027 was $3,735 million, which was 3.3 times the baseline. This aggressive scenario assumed screening adults four times annually and required an average of 228 million door-to-door symptom screenings and 15 million Xpert tests per annum (S Figure1). A large portion of the total cost was attributed to symptom screening (47%), followed by testing (25%) and treatment costs (8% for DS-TB and 7% for DR-TB).

In Max scenario 2, the estimated total budget for 2023-2027 was $13,438. A significant proportion of the cost was attributed to digital chest X-ray screening (75%) and Xpert testing (14%). In this scenario, 57 million digital chest X-ray screenings in a door-to-door context and 26 million Xpert tests would be implemented annually (S Figure1).

### Cost-effectiveness analysis

When estimating cost-effectiveness as the incremental cost per life year saved (LYS), among the individual NSP interventions, Xpert testing in symptomatic individuals seeking care at public health facilities (improving LYS by 14%) and providing TB preventive therapy for PLHIV (improving LYS by 1.1%) were cost-saving. Symptom screening for household contacts of patients with TB ($12/LYS), improving linkage to treatment ($13/LYS), and TUTT for individuals who are contacts of patients with TB ($84/LYS) were among the cost-effective interventions (Table 3). The packages of interventions included in the NSP scenario, and Max Scenarios 1 and 2, would cost $308/LYS, $712/LYS, and $3,774/LYS, respectively. A similar pattern emerged for incremental cost-effectiveness estimated as incremental cost per new active TB case averted and per DALY averted (S Tables 2 and 3).

**Table 3.**
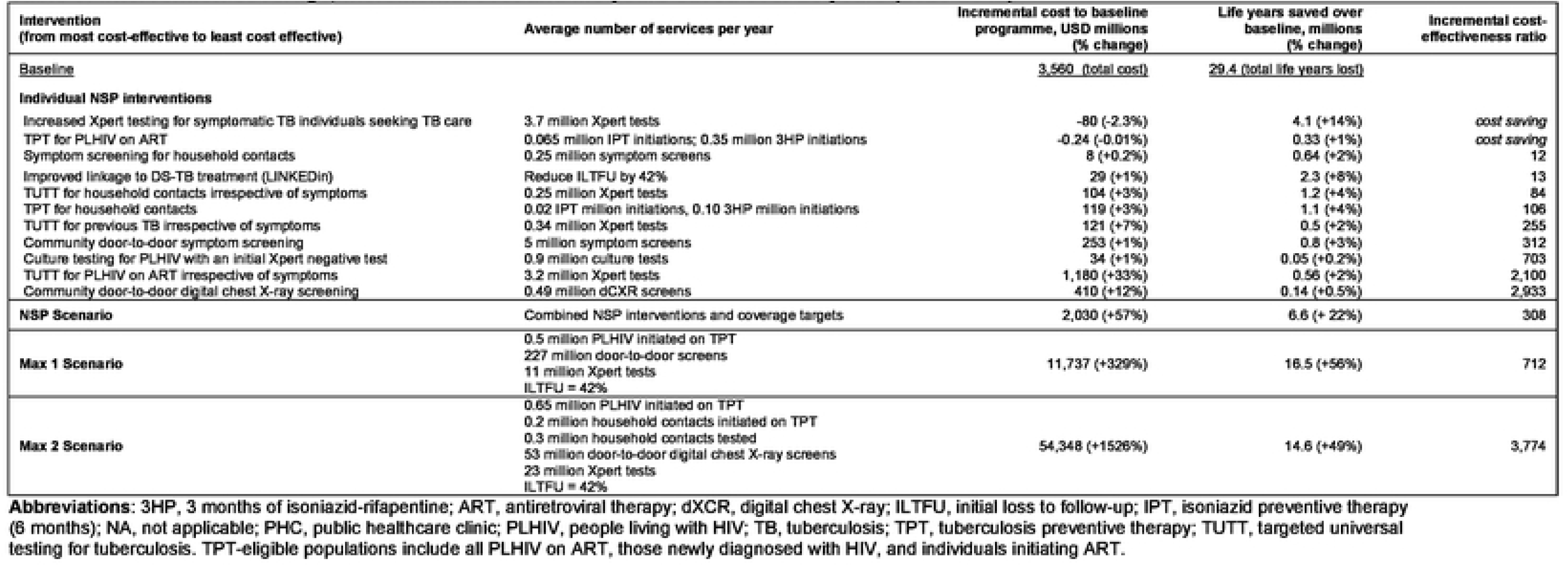
Intervention coverage. incremental cost and life years saved over 20 years (2023-2042)

## Discussion

We assessed the health impact and required budget for the TB component of South Africa’s National Strategic Plan for HIV, TB and STIs 2023–2027. We also evaluated the cost-effectiveness of individual and combined TB interventions in various scenarios over 20 years. Our analysis suggested that under the NSP, the most cost-effective interventions were increasing Xpert testing rates among individuals seeking TB care at health facilities, TPT for PLHIV and household contacts, increasing linkage to treatment, and universal testing for household contacts of TB patients. However, the NSP and the other more ambitious scenarios, while leading to significant reductions in TB incidence and mortality, fell far short of meeting the 2030 End TB targets.

On average, the estimated annual budget required for the NSP (2023-2027) was $256 million, with a major allocation to symptom screening and diagnostics (62%), followed by DS-TB treatment (18%) and DS-TB treatment (16%). This reflects South Africa’s focus on finding missing TB patients, as detailed in the National TB programme strategic plans [39]. In comparison, the 2019/2020 National AIDS Spending Assessment Plus report for TB prevention and care estimated 63% of funds were allocated to treating DR-TB, 20% to DS-TB treatment, and 16% to symptom screening and diagnostics [7]. These differences in cost distribution can be attributed to factors such as differing cost estimation methods and the longer duration of older regimens for DR-TB treatment and inpatient care, which incur higher costs. Nonetheless, our analysis also highlights the significant budget requirement for DR-TB treatment, considering the relatively small absolute number of DR-TB patients (4%).

Among the individual NSP interventions, increasing Xpert testing rates among people seeking treatment for TB symptoms at health facilities is both highly effective and cost-saving. This intervention addresses a critical gap, as testing rates in eligible individuals attending health facilities have historically been suboptimal in South Africa. Previous studies have reported that among individuals presenting with TB-suggestive symptoms, the proportion receiving laboratory tests ranged between only 3% and 49% [40–42]. Given this substantial under-testing of eligible patients, interventions targeting this gap have considerable potential to improve case detection while remaining cost-saving, as they primarily improve coverage of an existing, cost-effective diagnostic service. Our results also indicate that symptom screening and TUTT for household contacts of TB patients would be cost-effective. This is likely due to the relatively higher yield of identifying TB cases in this high-risk group compared with passive screening strategies [43], which explains the estimated cost-effectiveness, aligning with findings from other studies [44–46].

The individual NSP interventions for community screening, including door-to-door symptom checks and digital chest X-rays, had a modest impact on TB incidence and mortality and were less cost-effective than other interventions. These interventions reached only 5% of the adult population annually (per NSP-specified coverages) and were not targeted at specific risk groups. The high unit cost of digital chest X-ray combined with this low coverage may explain the limited cost-effectiveness. While active community-wide screening compared to passive screening has shown substantial impact in reducing prevalent TB in settings such as Vietnam [47,48], and the WHO endorses digital chest X-ray as a useful tool for detecting asymptomatic TB [49], cost remains a barrier to large-scale implementation. Alternative approaches that may improve cost-effectiveness would require less costly screening tools, targeting higher-risk populations or regions, for population-level scale-up.

Among the TUTT interventions, increasing TUTT in PLHIV was the least cost-effective strategy. The relatively low impact of this intervention on incidence and mortality may be a result of a saturation effect (existing screening interventions already targeting the PLHIV population). In addition, empirical studies showed that the yield of a positive TB diagnosis in PLHIV (4%) is much lower than in those with previous TB (11%) and household contacts (7%) [20,21]. This low yield, combined with the much larger target population of PLHIV (approximately 3 million compared to 0.3–0.4 million for household contacts and those with prior TB episodes), which drives up screening costs, explains the lower cost-effectiveness in this group. This highlights the need for identifying and targeting other population groups at risk of TB, particularly among people without HIV [26,50].

Our model indicated that providing PLHIV with TPT would be cost-saving, though it would have a modest impact on reducing TB incidence and mortality (<2%). Providing household contacts of TB patients with TPT was also among the most cost-effective interventions estimated to prevent 3% of new TB cases and reduce TB deaths by 4% between 2023 and 2042, for $106/LYS. In these TPT scenarios, we assumed 3HP would be scaled up to 90% coverage, and IPT would be maintained at 10%, from 2027 to 2042. Thus, the estimated impacts on incidence and mortality and cost-effectiveness likely reflect the inclusion and high coverage of 3HP (90%) and the assumption that its efficacy in curing infection in 50% and therefore longer-term reduction in TB disease risks [51,52]. Sensitivity analysis showed that the cost-effectiveness of 3HP was highly sensitive to assumptions regarding its sterilizing effect, with higher efficacy leading to greater health gains and improved cost-effectiveness. Our findings are nonetheless consistent with other studies suggesting that scaling-up preventive therapy could be cost-effective [53–55]. However, TPT implementation in South Africa currently falls short of targets for the eligible population.

The packaged interventions under the NSP scenario were estimated to have a substantial impact, with a 46% drop in incidence and a 48% reduction in mortality by 2030 compared to 2015, for $308/LYS over the period 2023-2042. Max Scenario 1, involving door-to-door symptom screening of all adults four times annually, further reduced incidence by 58% and mortality by 72%. Although total costs were three times higher than the baseline, they declined slightly over time (3–5%) as earlier diagnosis reduced transmission and prevented new cases (Supplementary Figures 3e and 3f). Max Scenario 2 included annual door-to-door digital chest X-ray screening of all adults. This more sensitive screening approach achieved a similar impact (56% reduction in incidence, 68% in mortality), but increased costs considerably (by 1526%), with chest X-rays accounting for 75% of total spending. These hypothetical scenarios are unlikely to be feasible as they require significantly expanded laboratory capacity and additional human resources, but have been included to illustrate the upper limits on what can be achieved with community-based screening.

**Figure 3:**
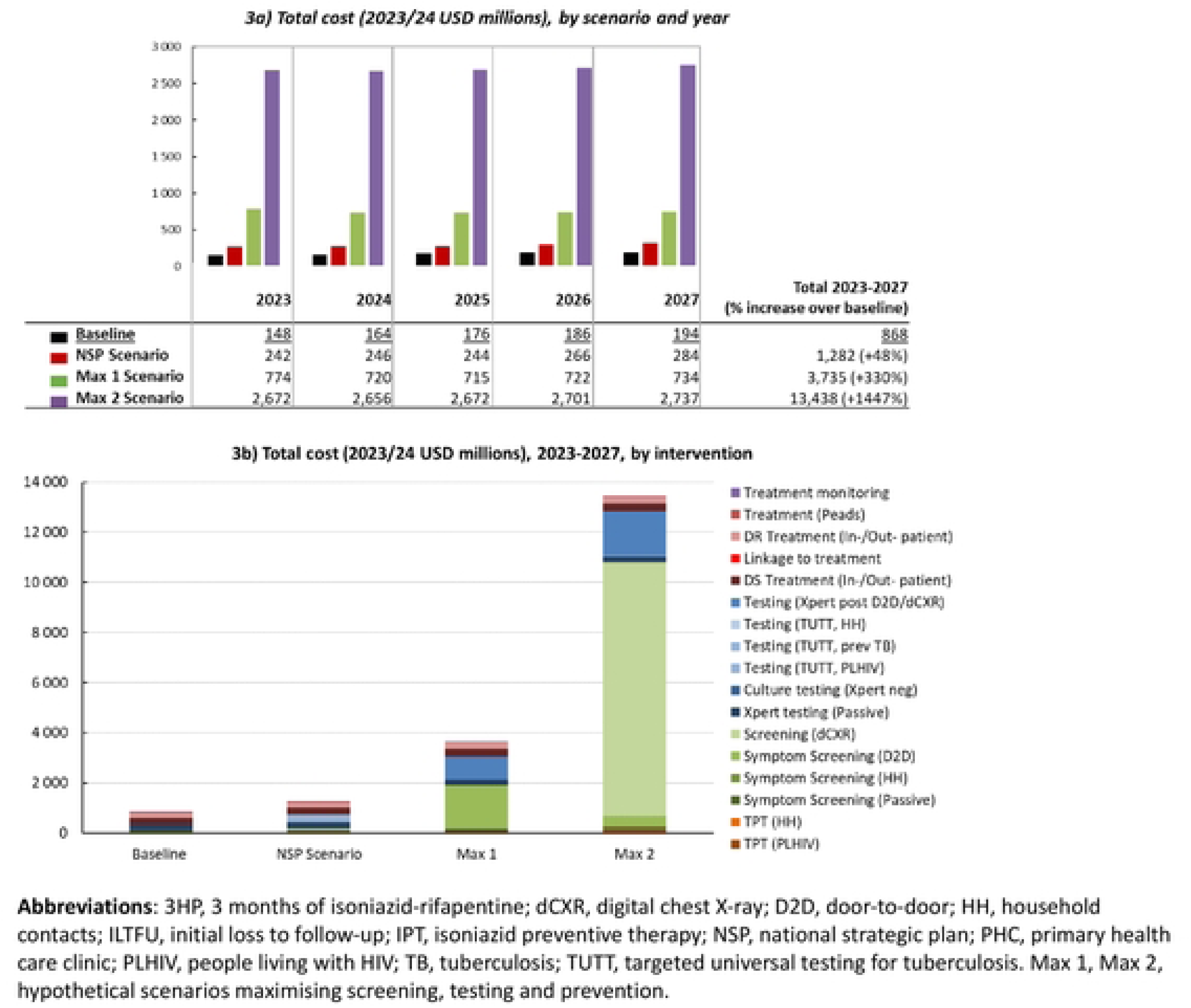
Estimated required budget 2023-2027.

Despite substantial reductions in TB incidence and mortality under these intervention scenarios, our model suggests that South Africa will not meet the End TB milestones of an 80% reduction in TB incidence and a 90% reduction in mortality by 2030 relative to 2015. While global and South African TB trends have been declining, the COVID-19 pandemic has disrupted TB control efforts [11]. However, even before the COVID-19 pandemic, it was noted that progress towards reaching the End TB milestones had been slow [56]. The South African TB programme has implemented plans to recover from the effects of the COVID-19 pandemic [57], but meeting the End TB milestones will still be challenging.

Our analysis has several limitations. First, we did not consider costs from the patient’s perspective, as the aim of the analysis was to inform government spending. However, patient costs are important for service uptake and sustainability, and eliminating catastrophic costs experienced by TB patients is part of the End TB strategy. Second, our definition of TUTT in persons with a history of TB differs from that of the empirical studies; our analysis includes anyone who has ever had TB, whereas the NSP refers to individuals who have had TB in the last two years. As a result, the less specific definition may underestimate the impact of TUTT in this TB risk group, given that the excess risk of TB in previously-treated TB patients is highest in the year after treatment completion [58]. Third, our modelling approach lacks a dynamic consideration of the paediatric population. Instead, we relied on 2023 WHO estimates to paediatric TB treatment, which accounts for 2% of the total cost. Therefore, this assumption does not accurately account for changes in the child population over time, nor does it link the incidence of TB in children to that in adults.

Additionally, the community-based screening interventions were not targeted to specific populations, which may have reduced their yield and cost-effectiveness. Our analysis also did not model any recently developed interventions to address other TB risk factors (i.e., undernutrition, diabetes, alcohol use, and smoking), which contribute significantly to TB incidence and mortality [1,8]. We also did not test the impact of specifically targeting important high-risk populations such as miners, healthcare workers, correctional services officers, and incarcerated individuals. Lastly, we did not conduct a multivariable sensitivity analysis of the epidemiological model and intervention parameters; however, a related analysis using the same model assessed the interventions most critical to achieving the End TB targets in South Africa [30]. Consistent with our findings, the analysis showed that TB testing for individuals seeking care at public health facilities could lead to substantial reductions in TB incidence and mortality. The study also highlighted the need for interventions targeting modifiable risk factors such as diabetes and smoking [30]. Altogether, within the context of these limitations and specified model assumptions, these projections, while not definitive, can indicate which aspects of the current TB programme require the most attention to achieve significant future TB reductions.

Our analysis suggests that the NSP packaged interventions would lead to substantial declines in TB incidence and mortality; however, much greater declines would be required to meet the End TB milestones. To accelerate progress, remaining priorities include accelerating the development of TB vaccine development, low-cost diagnostic tools, and shorter TB treatment regimens, while targeted interventions for hitherto unreached population groups (including men) to improve their access and engagement in care, and addressing broader determinants of TB, such as undernutrition, diabetes and socioeconomic barriers, are also crucial.

## Data Availability

All relevant data are within the manuscript and its Supporting Information files.

